# Clinic-Based Prevalence of Speech and Language Disorders Among Preschoolers: Implications for Pediatric Developmental Screening in Ghana

**DOI:** 10.1101/2025.11.11.25339988

**Authors:** Benedict Marfo, Emmanuel Kwaku Addo

**Author notes:** **Author 1** Benedict Marfo, Department of Community Health, University of Ghana Medical School, College of Health Sciences, University of Ghana, Accra, Ghana; **Author 2** Emmanuel Kwaku, Addo, Binghamton University, 4400 Vestal Parkway East, Vestal, NY 13850, USA, Department of Community Research and Action Binghamton, USA.

## Abstract

**Purpose:** This exploratory study examines the clinic-based prevalence and types of speech and language challenges among preschoolers referred by physicians to the speech-language therapy clinic of the Korle-Bu Teaching Hospital (KBTH) in Accra. It also explores the associations between preschoolers’ gender, age, and stuttering and highlights implications for pediatric development care.

**Method:** This retrospective descriptive study reviewed clinical records of preschoolers referred to the speech and language therapy unit of the KBTH. Data were extracted on age, gender, diagnosis, and referral source. Descriptive statistics were used to determine the prevalence and distribution of speech and language difficulties, and associations between demographic characteristics and disorder type were explored.

**Results:** Of the children referred, the majority were males, with a male-to-female ratio of approximately 3:1. The most common diagnoses were speech and language delay (87.7%), followed by stuttering (4.5%), receptive language disorder (2.6%), expressive language disorder (1.9%), articulation disorders (1.3%), and speech sound disorders (0.6%). Developmental delay was the most common medical diagnosis (48.7%). A substantial proportion of cases involved children aged 2–3 years, indicating referrals within the critical developmental window for early intervention.

**Conclusion:** Speech and language delay was the most common communication disorder, while developmental delay was the most frequent medical condition among preschoolers referred for therapy. Findings highlight the need for increased awareness among physicians and primary care providers regarding early communication screening and referral. Strengthening multidisciplinary collaboration and integrating speech-language assessment into routine child health services could enhance early detection and intervention for communication disorders in Ghana and similar low-resource contexts.

---

Speech and language disorders are among the most common developmental challenges in early childhood, often arising from complex or multifactorial causes (Tchoungui Oyono et al., 2018). While some cases are linked to identifiable medical conditions such as neurological disorders or developmental syndromes, many remain idiopathic. These disorders not only impair communication but may also serve as early markers of broader neurodevelopmental conditions (Rosenbaum & Simon, 2016). The consequences of speech and language disorders are far-reaching. When left unaddressed, they can disrupt academic progress, emotional well-being, social inclusion, and quality of life, with effects that persist into adulthood (Addo et al., 2025; Feeney et al., 2012; Langbecker et al., 2020; Mrini et al., 2025). As a result of these challenges, adults with a history of childhood communication disorders are at greater risk of underemployment and reduced educational attainment (Johnson et al., 2010; Law et al., 2009). Given these lifelong impacts, early detection and intervention are crucial for improving outcomes.

Globally, clinic-based studies consistently identify language delay as the most common speech and language disorder among referred children, with higher prevalence rates often reported in males (Gharib et al., 2017; Memon et al., 2024). These studies provide vital insight into the burden of communication disabilities and highlight the need for early, targeted intervention. However, most evidence comes from high-income or middle-income contexts, and patterns from low-resource settings remain underexplored.

Despite advances in speech-language pathology (SLP) training and healthcare infrastructure, a significant evidence gap remains in Ghana. No study to date has documented the clinic-based prevalence or patterns of speech and language disorders among Ghanaian preschoolers, despite the country’s rich linguistic diversity and growing rehabilitation services. This lack of empirical data may limit stakeholder efforts to design culturally appropriate assessment measures, evidence-based policies for early referral among caregivers, and programs for early childhood communication development.

To address this gap, the present study provides the first clinic-based prevalence estimates of speech and language disorders among preschoolers referred to the Speech Therapy Clinic at a university-affiliated hospital between 2019 and 2021. It also examines referral patterns by age, gender, and comorbid diagnoses. By generating baseline data from this hospital, this study offers novel insight critical for guiding policy, informing local training priorities, and strengthening service delivery.

## Prevalence of Speech and Language Disorders in Children

The prevalence of speech and language disorders in children varies across regions and is influenced by factors such as study methodology, stuttering identification methods, participants’ age, and other demographic differences (McCormak et al., 2007). The communication disorders literature is replete with prevalence data collected from both clinical populations and the general population. A review of data from both clinical and general populations shows that language delays are among the top diagnoses among children (Memon et al., 2024; Rezaei & Talimkhani, 2025). For example, a cross-sectional study conducted at a pediatric rehabilitation ward in Pakistan found that language delay was the most common disorder, affecting 61.8% of children aged 2 to 10 years (Memon et al., 2024). The study also found that males were disproportionately affected, and a significant proportion of children had a family history of speech-language disorders. Socioeconomic status also played a role, with the majority of affected children coming from middle-class families. Additionally, a recent study from Iran by Rezaei and Talimkhani (2025) analyzed 1,468 hospital records of patients referred to speech therapy clinics in Iran between 2013 and 2022. Their findings indicate a higher burden of communication disorders in children aged three to six, with speech-sound disorders accounting for 18.9% of cases, followed by speech and language delay (18.1%), stuttering (18%), voice disorders (12.1%), developmental language disorders (9.2%), and swallowing disorders (2.5%). However, despite the valuable contribution of their findings, Rezaei and Talimkhani did not provide sufficient discussion to contextualize their prevalence estimates. For example, although they reported prevalence data from hospital-based samples, the distinction between clinic-based and population-based estimates was not clearly articulated in their discussions. This omission may inadvertently contribute to challenges in accurately interpreting the true population burden of speech and language disorders. It is noteworthy that, given the prior identification of developmental concerns by caregivers and healthcare professionals that prompted referral for further assessment, prevalence estimates in this clinical population are expected to be higher than those reported in population-based studies.

In contrast to data from clinical samples, population-based prevalence studies are typically conducted through community or school-based screenings, aiming to provide accurate estimates of the true burden of communication disorders in the general population (McLeod & McKinnon, 2007). These studies tend to report comparatively lower prevalence rates but offer valuable insights into broader trends and service gaps (McCormack et al., 2007). A study in rural South India found an overall communication disorder prevalence of 4.29% among school-aged children, with 1.04% presenting with speech and language disorders and a larger proportion affected by hearing-related issues (Ravi et al., 2021). Additionally, McKinnon et al. (2007) conducted a large-scale study of 10,425 primary school students in Australia, identifying speech-sound disorders and stuttering as the most frequently diagnosed communication disorders, with prevalence rates of 1.06% and 0.33%, respectively. Voice disorders were also reported, though at a lower rate of 0.12%.

Despite the growing global interest in early childhood communication disorders, prevalence data on speech and language disorders in children from Africa remain scarce. Within the past decade, the few studies conducted in African countries to examine the prevalence of diagnosed speech and language disorders among preschoolers have found varying and incomparable reports. For instance, a population-based study conducted in Yaoundé, Cameroon, by Tchoungui Oyono et al. (2018) assessed 460 children aged three to five across seven communities and reported a stuttering prevalence of 8.4%, articulation and voice disorders at 3.6% each, receptive language disorders at 3%, and expressive language disorders at 1.3%. An earlier hospital-based study conducted in Egypt found that 10% of children aged three to six years attending an outpatient clinic had a confirmed diagnosis of communication disorders (Gharib et al., 2017). According to the team, delayed language development accounted for 6.4% of the cases, speech disorders for 3.2%, and voice disorders for 0.4%. The study also identified contributing factors such as sex, perinatal or postnatal complications, and existing medical conditions. These results suggest that, although little is known about the burden of speech and language challenges among children in Africa, it is important that research in this area is increased to identify the specific needs of this population.

## Diagnosis and Referral Patterns in Pediatric Speech and Language Disorders

Chen et al. (2022) have suggested that neonatal and maternal health conditions may contribute to communication challenges in young children. However, early identification before children start school has been shown to improve early interventions and improve school outcomes (Wallace et al., 2015). Pediatric health care providers often serve as the first point of contact for concerned caregivers and play a critical role in identifying early signs of speech and language disorders during routine health visits (Cohen et al., 1998). As primary caregivers who spend lots of time with their children, parental reports of communication challenges are often overlooked by pediatricians in their decision-making process for referrals to SLPs (Winters & Byrd, 2020). Instead, the pediatricians base their referral decisions on observable speech behaviors or maladaptive communication attitudes of these children during visits. The potential flaw in this approach is that some children can fall through the cracks and not receive the care that they need if they do not show these maladaptive behaviors or communication attitudes during their hospital visits. This could lead to pediatricians or other health workers advising parents to adopt a wait-and-see approach. Byrd and Winters (2020) contend that although the tendency to adopt a “wait and see” approach has decreased among pediatricians in the US in their referral of children who stutter for SLP follow-up, the absence of standardized referral criteria may lead to variability in identification and under-referral of children who require early intervention. Additionally, although early observations and parent reports play an important role in recognizing developmental issues, the formal assessment and treatment by SLPs are central, as their expertise is strongly linked to better language development outcomes (Roulstone et al., 2003). Importantly, it is worth stating that the initial diagnoses or suspicions raised by physicians may not always align with the refined diagnostic conclusions reached by SLPs following comprehensive evaluations.

Diagnosing speech and language disorders is inherently complex and shaped by sociocultural contexts. This is exacerbated by the variations in communication norms, which influence how physicians perceive developmental concerns and determine the urgency of referrals (Damico et al., 2021). Interprofessional collaboration between physicians or healthcare professionals and SLPs is therefore essential, as it facilitates accurate, timely diagnoses and supports the development of individualized interventions. Such partnerships have been shown to enhance outcomes for children with communication disorders. Despite the recognized benefits of interprofessional collaboration between physicians and speech-language pathologists (SLPs), there is a lack of research in Ghana that documents the specific conditions for which physicians refer preschoolers to speech therapy clinics. As Rezaei and Talimkhani (2025) note, understanding referral patterns and diagnostic outcomes is crucial for developing effective and targeted intervention strategies. At the university-affiliated hospital in Accra, pediatricians frequently refer children to the speech therapy clinic; however, the nature of referred disorders and the degree of alignment between initial medical diagnoses and subsequent SLP assessments remain underexplored.

## Methods

### Study design

This retrospective study involved reviewing SLP caseloads of preschoolers referred for therapy sessions at the speech-language therapy clinic of the Korle-Bu Teaching Hospital (KBTH), Accra, Ghana. The retrospective design enabled the examination of existing clinical records, providing insights into the types of speech and language disorders referred, their prevalence, and their demographic distribution. As the data reflect only preschoolers referred for evaluation, the findings represent a clinic-based prevalence of speech and language disorders within this referred population.

### Study Area

The study was conducted at the KBTH, which has a large bed capacity. The hospital comprises multiple clinical and diagnostic departments, including several specialized centers. The speech therapy clinic is located within the Ear, Nose, and Throat (ENT) unit. As of 2023, the clinic was staffed with six SLPs, and its caseload averaged 20 clients per week, totaling approximately 80 clients each month.

After receiving approval from the Community Health Department Review Committee of the University of Ghana Medical School (UGMS-CHDRC/120/2023) and consent from the hospital and its Ear, Nose, and Throat Unit, data were extracted from speech-language pathology clinical records. To protect confidentiality, all documents were de-identified before extraction, with names and hospital numbers replaced by unique codes. Only aggregated data were analyzed, and access to the original records was restricted to the research team. Electronic files were stored securely on encrypted, password-protected institutional servers. The study was conducted in accordance with Ghana’s Data Protection Act (2012) and the Declaration of Helsinki. The dataset comprised records of preschoolers who attended therapy sessions between 2019 and 2021. Information collected included demographic details, medical diagnoses provided by referring physicians, and SLP clinical diagnoses related to speech and language disorders.

### Study Population

The study population consisted of preschoolers, aged two to five years, referred by physicians for speech therapy services at a university-affiliated hospital. These preschoolers were seeking assistance for a variety of speech and language issues, including, but not limited to, stuttering, speech-sound disorders, and voice disorders. The inclusion criteria for data extraction were referrals of children aged 2 to 5 who received speech therapy services between 2019 and 2021, with their age, sex, and medical diagnosis documented. This ensured that only complete data sets were used in the analysis, eliminating potential bias due to missing demographic information.

### Data Analysis

The collected data were entered into Microsoft Excel (version 16.0) for initial processing and analyzed using the Statistical Package for Social Sciences (SPSS) version 20. All categorical variables were summarized as proportions. As an exploratory study, descriptive statistics were used to identify patterns. Of the 154 participants, some demographic information (gender and age) was missing at random; therefore, those cases were excluded, resulting in a subset of 118 participants for Fisher’s exact test analysis. Fisher’s exact test was conducted to examine the relationship between participants’ age, gender, and the prevalence of stuttering. This test was selected over the Chi-square test due to the expected frequencies of some cells being less than five (Kim, 2017). A *p*-value of less than 0.05 was considered statistically significant, indicating that the observed associations were unlikely to have occurred by chance.

## Results

A total of 154 preschool children, aged between 2 and 5 years, were included in this study. The majority were male, comprising 118 (76.6%), while females made up 36 (23.4%), as detailed in Table 1. Children aged 2–3 years constituted the largest proportion of the sample (77.9%), while those aged 4 to 5 constituted 22.1%. The mean age of participants was 3.4 years (± 0.8 SD). Analyses of gender and age associations with stuttering were conducted on 118 participants who had complete demographic data.

**Table 1.** Table Showing Demographic Statistics.

| Variable | Frequency | Percentage (%) |
| --- | --- | --- |
| Gender |  |  |
| Male | 36 | 23.4 |
| Female | 118 | 76.6 |
| Age (years) |  |  |
| 2-3 | 120 | 77.9 |
| 4-5 | 34 | 22.1 |

*Note.* The mean age of the participants is 3.4 years ( $\pm 0.8$ SD)

### Medical Diagnosis of Cases Referred

Among the participants, 75 (48.7%) were diagnosed with developmental delay. One-third (33.1%) of preschoolers were referred with a diagnosis of autism spectrum disorder (ASD), while 11 (7.1%) had cerebral palsy. Less common diagnoses included Down syndrome (1.9%), cleft palate (1.3%), seizure disorders (1.9%), and microcephaly (1.3%) (see Table 2 for details).

**Table 2.** Medical Diagnosis of Participants.

| Diagnosis | Frequency | Percentage (%) |
| --- | --- | --- |
| Developmental Delay | 75 | 48.7 |
| Autism Spectrum Disorder | 51 | 33.1 |
| Cerebral Palsy | 11 | 7.1 |
| Down Syndrome | 3 | 1.9 |
| Cleft Palate | 2 | 1.3 |
| Microcephaly | 2 | 1.3 |
| Seizure Disorder | 3 | 1.9 |
| Sickle Cell Disease | 3 | 1.9 |
| ADHD | 1 | 0.6 |
| Atrial Septal Defect | 1 | 0.6 |
| Bilateral Facial Nerve Palsy | 1 | 0.6 |
| Cerebral Atrophy | 1 | 0.6 |

*Note.* Percentages are rounded to one decimal place and may not sum exactly to 100 due to rounding. ADHD = Attention-Deficit/Hyperactivity Disorder

### Speech-Language Pathologist Diagnosis

As seen in Figure 1, the most common SLP diagnosis was speech and language delay, affecting 135 (87.7%) of the preschoolers. Other diagnoses included stuttering (4.5%), language disorder (2.6%), expressive language disorder (1.9%), and speech sound disorder (0.6%).

**Figure 1.**
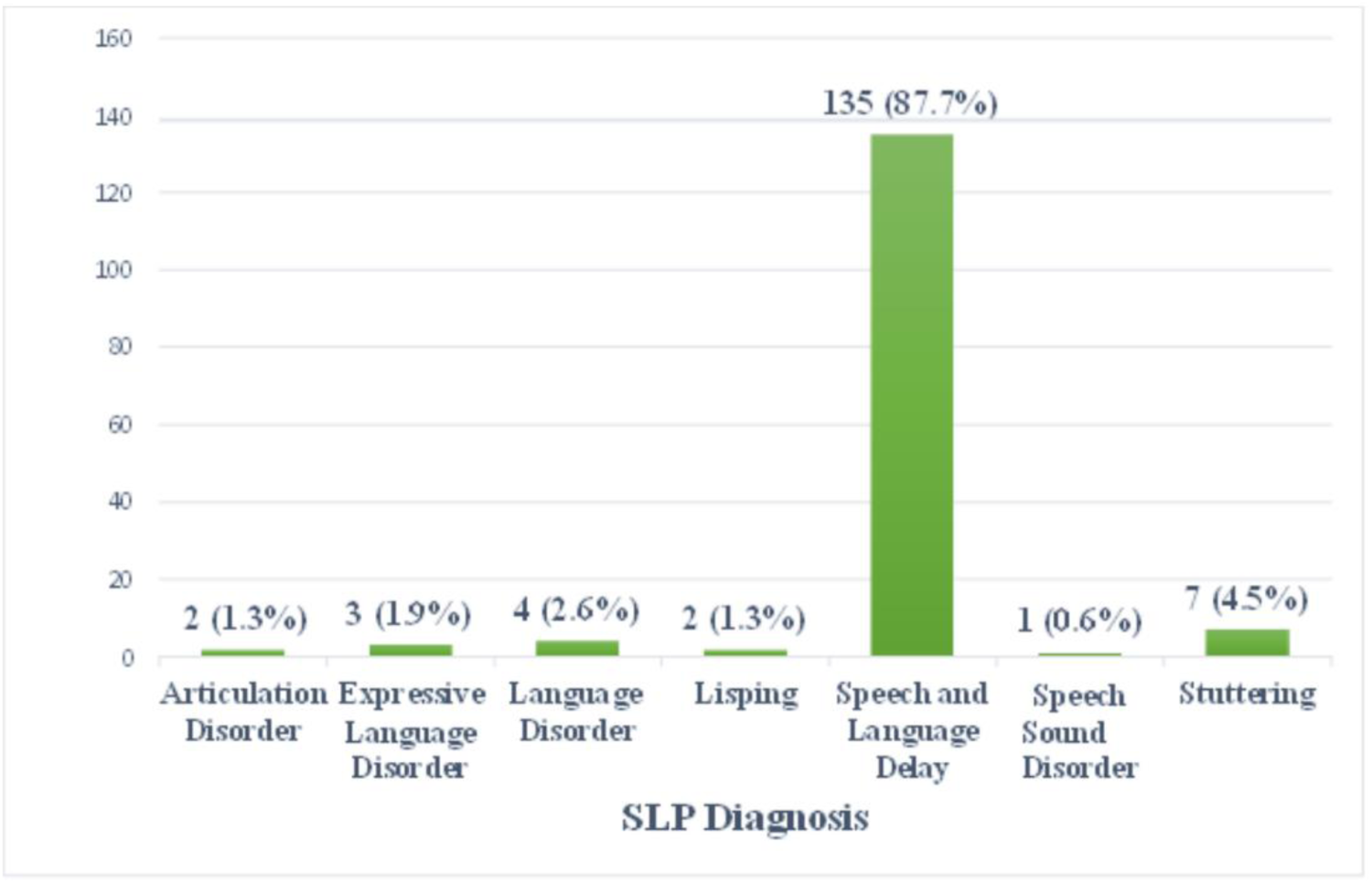
Distribution of SLP Diagnoses.

### Association between Stuttering, Age, and Gender

Fisher’s exact tests were conducted to examine demographic associations with stuttering. Among males, 5 of 82 (6.10%) were identified as stuttering, compared with 2 of 36 females (5.56%). The odds of stuttering for males were slightly higher than females (OR = 1.10, 95% CI [0.19, 6.24]), though this difference was not statistically significant (p = 1.000, φ = .02). When examined by age, 5 of 83 children aged 2–3 years (6.02%) and 2 of 35 children aged 4–5 years (5.71%) were reported to stutter. The odds of stuttering were similar between age groups (OR = 1.06, 95% CI [0.18, 6.31]), with no statistically significant association observed (p = 1.000, φ = .01). See Table 3.

**Table 3.** Fisher’s Exact Test of Association Between Gender, Age and Stuttering Among Preschoolers.

| Variable | Stuttering<br>n (%) | No Stuttering<br>n (%) | p-value | OR (95% CI) | $\phi$ |
| --- | --- | --- | --- | --- | --- |
| Gender |  |  |  |  |  |
| Male | 5 (6.10) | 77 (93.90) | 1.000 | 1.10 (0.19, 6.24) | .02 |
| Female | 2 (5.56) | 34 (94.44) |  |  |  |
| Age |  |  |  |  |  |
| 2-3 | 5 (6.02) | 78 (93.98) | 1.000 | 1.06 (0.18, 6.31) | .01 |
| 4-5 | 2 (5.71) | 33 (94.29) |  |  |  |

*Note.* Fisher's exact test used due to expected cell counts <5. Effect size estimates are reported using odds ratio (OR) and phi ( $\phi$ ). CI = confidence interval.

## Discussion

This study aimed to determine the clinic-based prevalence and patterns of speech and language disorders among preschoolers referred to the speech-language therapy clinic at the KBTH in Accra between 2019 and 2021. Our secondary aim of examining the association between age and gender among our sample showed no statistical significance. Further, the findings indicated that speech and language delay was the most prevalent SLP diagnosis, affecting 87.7% of the sample. This was followed by stuttering, observed in 4.5% of cases. Other diagnoses included receptive language disorders (2.6%), expressive language disorders (1.9%), articulation disorders (1.3%), lisping (1.3%), and speech sound disorders (0.6%).

### Clinic-Based Prevalence of Speech and Language Disorders

The predominance of speech and language delay in this study reflects a broader global pattern observed across clinical populations. Similar trends have been reported in pediatric rehabilitation settings in Pakistan (Memon et al., 2024), South Korea (Lee & Choi, 2024), and Iran (Mehri et al., 2016), where language delay consistently emerges as the most frequent diagnosis among children referred for speech-language services. There is variation in prevalence rates across studies. The lower figure reported by Gharib et al. (2017) in Egypt, may be attributed to differences in referral practices, service accessibility, or diagnostic criteria (Rezaei & Talimkhani, 2025). These findings collectively highlight the importance of strengthening early screening and intervention systems, particularly in contexts where delays in identification could exacerbate long-term developmental challenges.

Beyond language delay, this study’s findings regarding other communication disorders among referred preschoolers provide valuable insight into patterns of service utilization and potential gaps in identification and referral. The prevalence of stuttering observed in our clinical sample is lower than that reported in comparable studies from other regions. For example, Mehri et al. (2016) identified stuttering in 21.2% of referrals, whereas Rezaei and Talimkhani (2025) reported a rate of 18% among children referred to speech therapy clinics in Iran. These differences may reflect variations in parental concern, physician detection practices, or cultural perceptions of stuttering, which could influence the likelihood of referral during early childhood. It is possible that in our setting, stuttering in preschoolers is perceived as part of normal developmental disfluency, leading to healthcare professionals and caregivers adopting a waiting approach rather than early referral for specialist evaluation. Similarly, the low referral rates for articulation, speech-sound, and voice disorders contrast with figures from other clinic-based studies. Mehri et al. (2016) reported that 21.6% of their cases had articulation and voice disorders, while Rezaei and Talimkhani (2025) found speech-sound disorders in 18.9% of the referred children. This disparity may point to under-recognition of these disorders by caregivers or primary care providers in our context, or possibly to prioritization of more overt language delays in referral decisions. Cultural and linguistic factors may also contribute to the decision-making process regarding what is perceived as typical versus disordered speech and language among preschoolers (Ndung’u & Kinyua, 2009; Washington et al., 2023).

In addition, the low occurrence of specific receptive and expressive language disorders in our referrals may reflect diagnostic overlap, where these conditions are subsumed under broader speech and language delay diagnoses at the point of referral. This pattern calls for detailed diagnostic evaluation by SLPs within our context to inform specific diagnoses and guide tailored interventions.

### Underlying Medical Diagnosis

In terms of associated medical conditions, developmental delay emerged as the most frequent diagnosis among children referred for speech-language services in this study, followed by autism spectrum disorder, cerebral palsy, Down syndrome, and seizure disorders. This pattern is consistent with findings from other clinical settings. For example, Memon et al. (2024) also identified seizure disorders as a common medical comorbidity among children with speech and language difficulties in a pediatric rehabilitation context. The prevalence of neurodevelopmental and neurological conditions among referred children demonstrates the well-established connection between these diagnoses and communication challenges, underscoring the critical importance of interprofessional collaboration between primary healthcare staff and SLPs (Lubker, 1997; Tchoungui Oyono et al., 2018).

### Gender and Age Distribution

The current study revealed a pronounced male predominance (76.6%) among referrals for speech and language evaluations, consistent with existing literature documenting higher prevalence rates of speech and language disorders in males. For example, Addo et al. (2025) in their study among school-aged children reported that 87% of the children who stuttered were male. Aslam et al. (2020) and Ravi et al. (2021) similarly found boys to be disproportionately affected by speech sound disorders. These patterns likely reflect underlying biological, developmental, and socio-environmental factors contributing to males’ increased vulnerability, which may drive their overrepresentation in early intervention settings. Age distribution data highlighted the importance of early identification efforts, with most referrals occurring between two and three years of age. This aligns with global recommendations that emphasize early detection during periods of heightened neuroplasticity (McLaughlin, 2011; WHO, 2012). Strengthening referral pathways remains critical, particularly in Ghana, to optimize early intervention and reduce long-term communication impairments.

### Association Between Age, Gender, and Stuttering

Using Fisher’s exact test, this study found no statistically significant association between stuttering and either gender or age group, with negligible effect sizes among preschoolers. While this finding aligns with prior research indicating that gender and age-related disparities in stuttering prevalence often emerge more clearly after the preschool years (Manning & DiLollo, 2023; Reilly et al., 2009; Yairi & Ambrose, 2013), it contrasts with the findings of Mehri et al. (2015). Mehri and his colleagues reported significant gender differences in speech and language disorders among preschoolers. Despite the findings of this study extending the theory of a lack of disparity in stuttering prevalence among genders in preschoolers, there might be other reasons accounting for it. For instance, the absence of significant associations in our sample could be due to the limited number of stuttering cases (n = 7) identified. This could have reduced the statistical power of the data and increased the likelihood for Type II error (Charan & Biswas, 2013). With such small subgroup frequencies, it becomes difficult to detect subtle associations that may exist in the broader population. Though exploratory, our study provides data on early stuttering characteristics of Ghanaian preschoolers referred to the SLP clinic of a university-affiliated hospital. These findings help to address a critical gap in the literature and offer a valuable foundation for future research into the early identification of stuttering cases and physician referrals in low-resource settings.

### Implications for Practice and Policy

The findings from this exploratory study have significant implications for clinical practice, research, and health policy. The high prevalence of speech and language delay among preschoolers referred for speech and language therapy highlights the critical need for systematic early childhood communication screening within pediatric and primary care services. Continuous developmental assessment by child health and primary care practitioners should include communication milestones to ensure early identification and referral for early intervention.

From a clinical practice standpoint, the predominance of non-specific “speech and language delay” diagnoses points to the urgent need for standardized diagnostic protocols and locally normed assessment tools for the Ghanaian context. Developing culturally and linguistically appropriate screening and diagnostic instruments will enhance diagnostic specificity, support individualized treatment planning, and improve data comparability across diverse service settings (Washington et al., 2023). Also, SLP training should emphasize the importance of differential diagnosis and early caregiver education to strengthen clinical capacity within the Ghanaian context.

At the policy level, these findings highlight the need to enhance the integration of communication disorder screening within child health programs. Policymakers should consider embedding SLP services within maternal and child health frameworks, alongside nutrition, immunization, and early education initiatives, to ensure equitable access to early intervention. Strengthening interprofessional collaboration, particularly between pediatricians, SLPs, and early educators, should be prioritized through joint training programs and clear referral protocols.

Furthermore, the gender disparity observed in this study suggests potential sociocultural influences on health-seeking behaviors and caregiver perceptions of communication difficulties. National awareness campaigns emphasizing the importance of early speech and language development could help counteract stigma, promote caregiver vigilance, and encourage earlier engagement with SLP services.

Finally, this study provides a foundation for future large-scale population and clinic-based studies in other tertiary facilities and populations. This will provide data that could be used to inform policy development in Ghana. The Ministry of Health, in collaboration with the Ghana Health Service and health training institutions, could leverage these data to establish guidelines for developmental screening, referral pathways, and data reporting on communication disorders. Integration of SLP services into the National Health Insurance Scheme in Ghana could further expand accessibility for children from low-income households. Collectively, these measures would help strengthen Ghana’s early intervention infrastructure and improve communication outcomes for preschool-aged children nationwide.

### Limitations

This exploratory descriptive study had limitations that should be considered when interpreting the findings. First, as a clinic-based investigation conducted in one facility, the results may not be generalizable to the broader Ghanaian preschool population who seek treatment elsewhere. Children captured in this study are more likely to present with severe or complex communication disorders, as the sample context was a major teaching hospital in the capital city, Accra. This could inflate the clinic-based prevalence compared to community settings. To mitigate this, this study focused its interpretation on patterns of service utilization rather than population prevalence. Future studies should consider population-based studies to obtain a more representative sample that can be generalized.

A key strength of this study is its inclusion of a large, clinically referred sample of preschoolers, providing comprehensive insight into the prevalence of speech and language disorders in a hospital context. The dataset captured a broad range of diagnoses, and Fisher’s exact test was used to account for small subgroup sizes. However, the retrospective design limited control over data completeness and consistency, with missing gender and age information reducing the sample size for analyses of associations with stuttering. Consequently, findings regarding the relationship between age, gender, and stuttering should be interpreted with caution. Future studies should implement standardized data collection procedures to ensure complete demographic information, thereby enhancing the robustness of inferential analyses.

Third, while qualified SLPs made all diagnoses, the study did not document whether diagnostic tools were consistently applied. This may have introduced variability in diagnostic categorization. Nonetheless, assessments occurred within a specialized clinical setting, lending confidence to the professional validity of the diagnoses.

Despite these limitations, the study provides essential preliminary data on clinic-based prevalence and patterns of speech and language disorders in Ghanaian preschoolers, offering a foundation for future population-based and longitudinal research.

## Conclusion

This study provides valuable preliminary data on speech and language disorders among preschoolers referred to the KBTH in Accra, Ghana. The marked male predominance and early age of referral underscore critical windows for intervention, suggesting potential sociocultural influences on help-seeking behaviors. These findings highlight the continuous need to strengthen Ghana’s early childhood development framework by implementing enhanced screening protocols, improving referral pathways, and providing specialist training tailored to local linguistic and cultural contexts.

## Acknowledgements

Our heartfelt gratitude goes to Dr. Kenneth Klufio and Mr. George Aryee for their timely revisions and suggestions, as well as to Mrs. Malaika Sundiata and the entire team of speech-language therapists at the Korle-Bu Teaching Hospital for their support.

## Data availability statement

The data supporting this study’s findings are available on request from the corresponding author.

## Author contributions statement

B.M. conceptualized the study, collected the sample, conducted the analysis, and contributed to the writing/editing.

E.K.A. contributed to study design, analysis, writing/editing, and review.

## DECLARATIONS

### Ethics

Ethical approval with reference number UGMS-CHDRC/120/2023 was received from the Community Health Department Review Committee of the University of Ghana Medical School. The study was conducted in accordance with Ghana’s Data Protection Act (2012) and the Declaration of Helsinki.

### Consent

Permission to extract data from speech-language pathology clinical records was sought from KBTH and the Ear, Nose, and Throat (ENT) Unit.

### Pre-Print Disclaimer

This manuscript is a preprint and has not been peer-reviewed. The findings and conclusions should not be interpreted as final or conclusive and may change following peer review. Please contact the corresponding author for the most recent version.

